# Impact of a Statewide Hearing-Targeted Congenital Cytomegalovirus Screening Mandate: An 11-Year Interrupted Time Series Analysis

**DOI:** 10.1101/2025.08.31.25334783

**Authors:** Aanchal Wats, Barbara L. Araujo, Camila Aparicio-Llorente, Julia Ganem, Isabela O. Oliva, Jeffrey R. Gruen, Carlos R. Oliveira

## Abstract

**Objective:** Congenital cytomegalovirus (cCMV) is a leading cause of potentially treatable non-genetic hearing loss, but many cases are missed at birth. In 2016, Connecticut mandated hearing-targeted cCMV screening for all newborns who fail their initial hearing test. We evaluated its impact on diagnosis rates and long-term outcomes by comparing the 3 years of no screening with the subsequent eight years of mandated screening.

**Methods:** We conducted a retrospective interrupted time series study (2013–2023) within a health system serving ∼60% of Connecticut’s population. Confirmed cases were defined as positive CMV testing within 21 days of life. Annual diagnosis rates were modeled before and after mandate implementation. Long-term audiologic and neurodevelopmental outcomes were tracked through June 2025.

**Results:** Among 197,177 births, 48 infants had confirmed cCMV. The screening mandate was associated with a 3.6-fold increase in diagnoses (IRR 3.58; 95% CI 1.17–11.0), peaking at 4.09 per 10,000 births. Prior to the mandate, all detected cases were moderately to severely symptomatic. Afterward, 11 clinically inapparent cases were identified, 73% of whom were later found to have significant sequelae. Among those with sensorineural hearing loss at birth, 81% worsened over time. Developmental delays occurred in 61% of children with follow-up.

**Conclusion:** Hearing-targeted screening increased cCMV case detection and identified infants with clinically meaningful disease who would otherwise have been missed. However, many infections likely remain undetected, underscoring both the value and limitations of targeted approaches and highlighting the need to consider universal screening.

## Introduction

Cytomegalovirus (CMV) is one of the most common congenital infections worldwide, affecting 0.5%–0.7% of live births.^1,2^ In the United States alone, 20,000–30,000 infants are infected annually, of which more than 6,000 children develop CMV-related sequelae.^3–6^ Clinical manifestations of congenital CMV (cCMV) range from mild, non-specific findings, such as petechial rash, jaundice, and hepatosplenomegaly, to severe neurologic abnormalities, including microcephaly, sensorineural hearing loss (SNHL), and developmental delays. SNHL is the most common long-term sequela, affecting roughly 30% of infected infants, with 18%–27% of these cases presenting as delayed-onset hearing loss.^7–10^

For infants with moderate to severe symptomatic disease, initiating antiviral therapy within the first month of life can improve long-term hearing and neurodevelopmental outcomes.^11–14^ However, due to the often subtle or non-specific clinical presentation, many infants with clinically significant cCMV are not diagnosed in time for effective intervention.^15,16^ This diagnostic gap has prompted calls for policies that enable earlier detection and intervention.^17^

The optimal cCMV screening strategy remains a subject of ongoing debate. Discussions have largely centered on either universal screening of all newborns or targeted screening of high-risk infants, such as those who do not pass their initial hearing screen.^18^ Connecticut initially adopted the latter, and enacted legislation in 2016 requiring that all infants who fail their newborn hearing screen undergo CMV testing.^19^ However, data on the impact of this targeted screening policy on the rates of cCMV diagnosis and subsequent outcomes remain limited.^20^

The primary aim of this study was to estimate the impact of Connecticut’s 2016 hearing-targeted screening law on the rate of confirmed cCMV diagnosis. Our secondary aims were to compare the clinical severity of cCMV cases diagnosed before and after the law’s implementation and characterize the long-term audiologic and neurodevelopmental outcomes. We hypothesized that the law would increase the frequency of diagnosis and identify children with milder presentations by capturing infections that previously went undetected.

## Methods

### Study Design and Setting

We employed an interrupted time series (ITS) design to compare the rates of cCMV diagnoses before and after the implementation of hearing-targeted cCMV surveillance within a large healthcare delivery network in Connecticut. This quasi-experimental design allowed us to assess the impact of a population-level policy change by examining changes in trends over time. The study was conducted within the Yale New Haven Health System (YNHHS), a healthcare network that includes five hospital systems and over 130 ambulatory and urgent care sites, all connected through a single electronic health record (EHR) platform. The institutional review board (IRB) at the Yale School of Medicine approved the study and waived the requirement for informed consent (HIC:2000036550). This study follows the Strengthening the Reporting of Observational Studies in Epidemiology (STROBE) reporting guidelines for observational studies.

### Study Population

The source population included all patients who received care at YNHHS and were born within its primary service area between January 1, 2013, and December 31, 2023. The YNHHS primary service area is a 55-town region in southern Connecticut that includes 1.9 million people, representing ∼60% of the state’s population (Figure S1). Limiting inclusion to this geographic area ensured a well-defined catchment population and enabled calculation of population-based incidence rates. Relevant clinical and laboratory data were electronically extracted from the EHR of YNHHS patients who were born during the study period. This data included patient sociodemographics, birth history, clinical diagnoses (problem lists, encounter diagnoses), newborn hearing screening results, and laboratory test results.

### Case Ascertainment and Definition

Potential cCMV cases were identified through a combination of laboratory results and diagnostic codes. Laboratory criteria included the detection of CMV DNA using polymerase chain reaction (PCR) or other nucleic acid amplification techniques (NAAT) from urine, saliva, whole blood, or cerebrospinal fluid. International Classification of Diseases (ICD-9, ICD-10) and Systematized Nomenclature of Medicine Clinical Terms (SNOMED-CT) codes were also utilized as supplementary data sources for identifying potential cCMV cases (see Table S1). Medical records of all potential cases were manually reviewed to identify “confirmed cCMV”, which required a positive diagnostic test within 21 days of life. For these confirmed cases, a standardized form was used to collect data from encounter notes on initial clinical findings, treatments administered, and subsequent audiologic and developmental outcomes.

### Clinical Classification and Outcomes Assessment

Confirmed cCMV cases were classified using two approaches (Table S1). First, we categorized patients as “clinically apparent” or “clinically inapparent” cCMV based on physical findings observed during the initial newborn examination, prior to any laboratory or imaging evaluation. Infants with findings such as microcephaly, abnormal muscle tone, hepatomegaly, splenomegaly, petechiae, or small-for-gestational-age status were classified as clinically apparent; those without such findings were classified as clinically inapparent. The second approach assigned a disease severity category based on a complete diagnostic evaluation, using previously established definitions.^21^ Briefly, asymptomatic cCMV was defined as having a normal hearing test (based on diagnostic audiological evaluation) and no abnormalities on physical examination, neuroimaging, or laboratory tests. Infants whose only attributable clinical sign was abnormal hearing were classified as cCMV with isolated SNHL. Mildly symptomatic disease was defined as having two or fewer manifestations (e.g., thrombocytopenia, petechiae, or mild hepatitis). Moderately to severely symptomatic disease included cases with multiple (>2) clinical manifestations, or neurologic involvement (e.g., microcephaly, seizures, intracerebral calcifications), or severe organ dysfunction.

Long-term outcomes were followed through June 30, 2025. Neurodevelopmental outcomes, such as developmental delays and neurologic impairments, were identified from EHR problem lists, diagnostic codes, and provider notes. All available audiology reports were reviewed to document the presence, laterality, severity, and progression of SNHL. Hearing loss was classified as mild (21-40 dB), moderate (41-70 dB), or severe (>70 dB), based on thresholds in the more severely affected ear, using Auditory Brainstem Response (ABR) or the average of the hearing thresholds at four specific frequencies (0.5, 1, 2, and 4 kHz), as previously described.^22^ In instances where specific audiometric thresholds were not reported, the audiologist’s qualitative classification (i.e., mild, moderate, or severe) was used. For the subset of infants with a follow-up hearing evaluation after 12 months of age, we compared their baseline result (within the first 6 months of life) to their most recent evaluation to classify progression as improved, stable, or worsened. We also documented the need for hearing aids or cochlear implants.

### Screening Procedures

Before January 1, 2016, cCMV testing at YNHHS was performed at the discretion of individual healthcare providers. Following implementation of Connecticut’s hearing-targeted cCMV screening law, standardized protocols were adopted. In addition to testing based on clinical suspicion, all infants who failed the newborn hearing screen underwent cCMV testing using saliva or urine. A positive screen triggered a referral to an infectious disease specialist for a comprehensive diagnostic evaluation, which included confirmatory testing, bloodwork, a head ultrasound, and referrals for ophthalmologic and audiologic assessment. The complete screening algorithm is shown in Figure S2.

### Statistical Analysis

Patient characteristics were summarized using medians and interquartile ranges (IQR) for continuous variables and frequencies with percentages for categorical variables. Group comparisons were conducted using the Kruskal-Wallis test for continuous variables and Fisher’s exact test for categorical variables, as appropriate.

Because screening policy is expected to affect ascertainment rather than true infection prevalence, our primary outcome was the rate of documented diagnoses. To assess the impact of the hearing-targeted screening policy on annual cCMV diagnosis rates, we fit a Generalized Estimating Equations (GEE) model with a Poisson distribution, a log link function, and a first-order autoregressive correlation structure. The GEE model included an indicator for the post-policy period to estimate the immediate level change, and an interaction between time and the post-policy indicator to estimate the change in trend. Additional terms were included to capture analogous level and trend changes associated with the COVID-19 pandemic. The logarithm of annual births for the catchment area, derived from official town birth records, was included as an offset to directly model incidence rates. The model coefficients were exponentiated to produce incidence rate ratios (IRRs) with corresponding 95% confidence intervals (CIs). A two-sided p-value < 0.05 was considered statistically significant for all analyses. All analyses were conducted using StataMP version 18 (StataCorp LLC, College Station, TX).

## Results

### Study Population

Between January 1, 2013, and December 31, 2023, there were 197,177 live births in the catchment area. Seventy-two infants met preliminary criteria for cCMV, of whom 48 were confirmed and included in the analysis (Figure 1).

**Figure 1.**
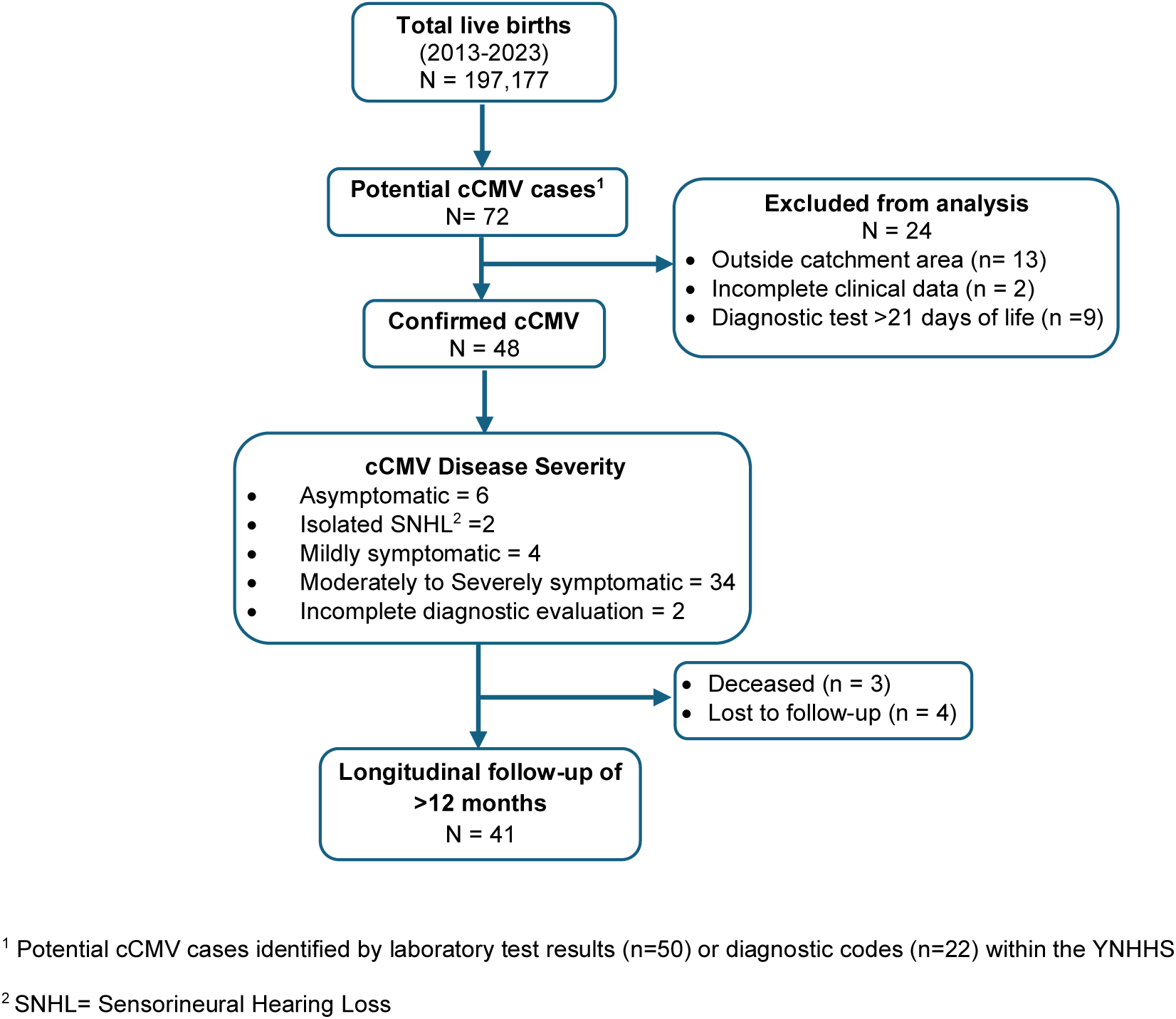
Flowchart of Study Participant Selection. Study participant selection showing 48 confirmed congenital cytomegalovirus (cCMV) cases from 197,177 live births (2013–2023), categorized as asymptomatic, isolated sensorineural hearing loss (SNHL), mildly symptomatic, or moderately to severely symptomatic.

### Clinical Characteristics and Long-Term Outcomes

The demographic and clinical characteristics of the 48 infants with confirmed cCMV are detailed in Table 1. The cohort was 46% female, and 48% were Non-Hispanic White. After a complete diagnostic evaluation, 34 (71%) were classified as moderately to severely symptomatic, 4 (8%) as mildly symptomatic, 2 (4%) as isolated SNHL, and 6 (13%) as asymptomatic cCMV (Figure 1). The most common findings at birth were small for gestational age or intrauterine growth restriction (24/48, 50%), microcephaly (14/48, 29%), abnormal muscle tone (10/48, 21%), and petechiae (8/48, 17%). Laboratory abnormalities included thrombocytopenia (18/48, 38%) and hepatitis (11/48, 23%). Abnormal neuroimaging was documented in 23/48 (48%), most often cysts (16/23, 70%), intracranial hemorrhage (8/23, 35%), or calcifications (4/23, 17%). SNHL at birth was documented in 16/48 (33%), with 9/16 (56%) having severe hearing loss. A total of 32/48 (67%) infants received antiviral therapy, of whom 25/32 (78%) completed the full course. Treatment-related toxicity occurred in 11/32 (34%), most commonly neutropenia (9/11, 82%).

**Table 1:**
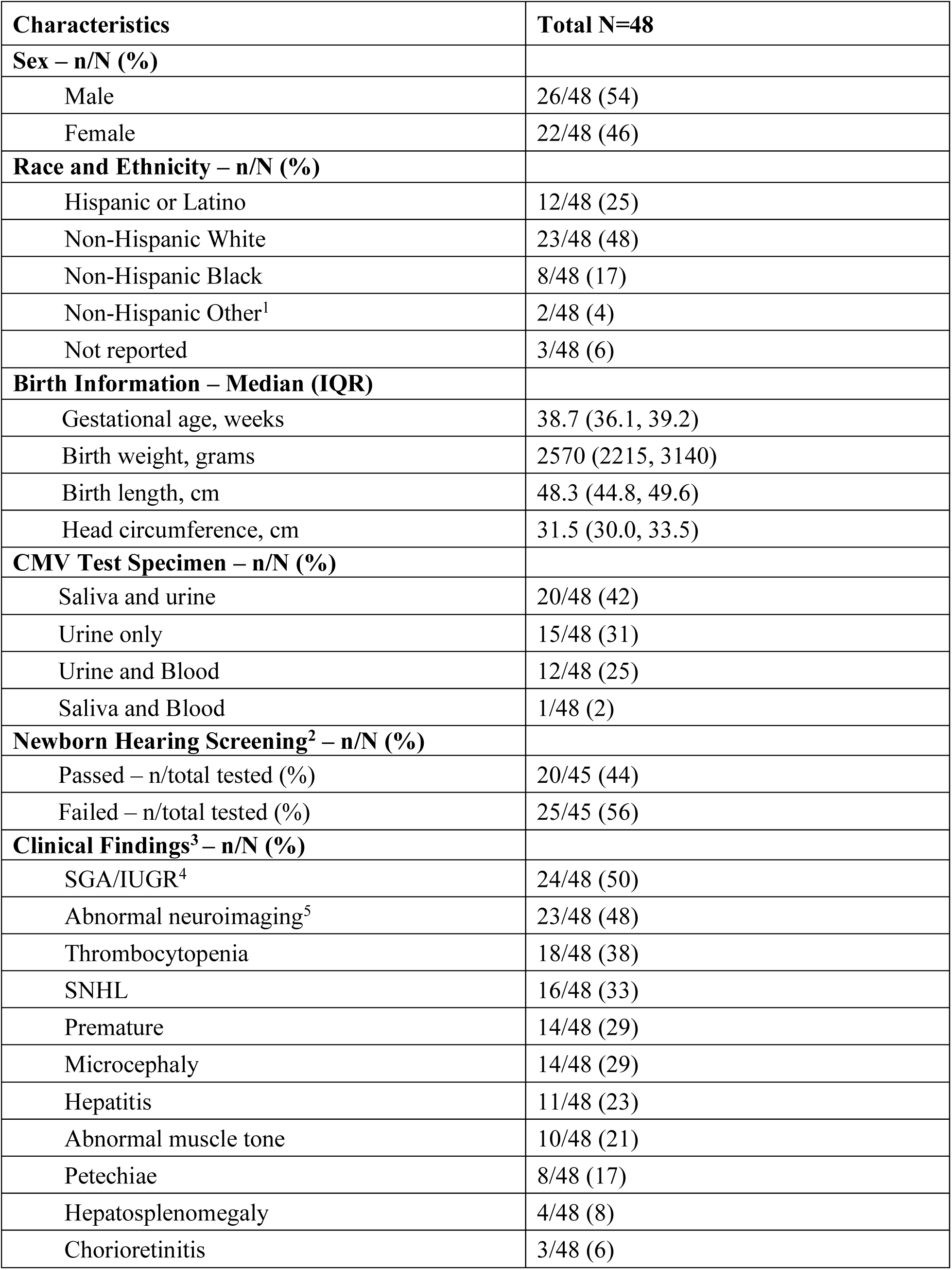
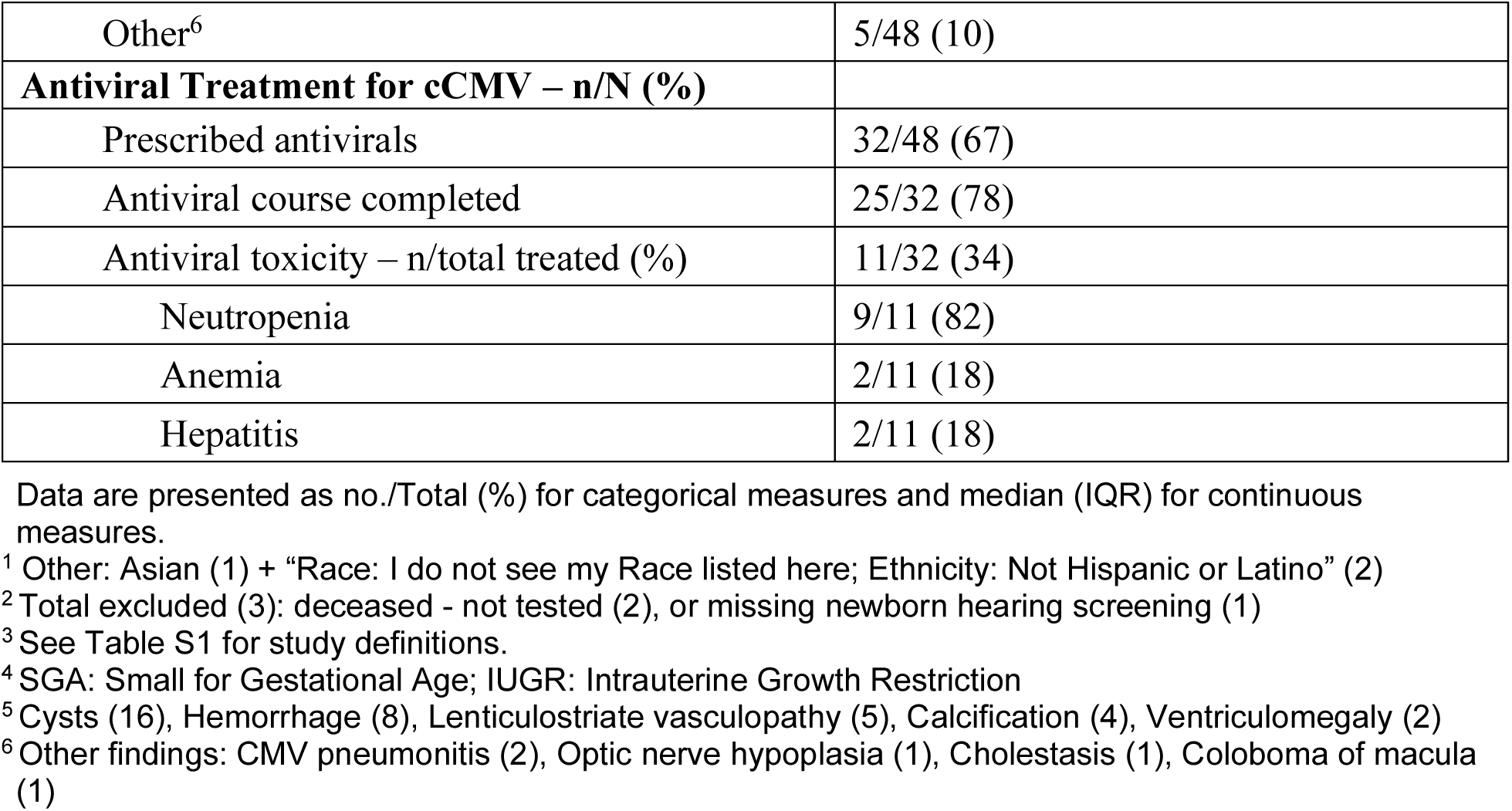
Characteristics of 48 Infants with confirmed cCMV Infection.

| **Characteristics** | **Total N=48** |
| --- | --- |
| **Sex – n/N (%)** |  |
| Male | 26/48 (54) |
| Female | 22/48 (46) |
| **Race and Ethnicity – n/N (%)** |  |
| Hispanic or Latino | 12/48 (25) |
| Non-Hispanic White | 23/48 (48) |
| Non-Hispanic Black | 8/48 (17) |
| Non-Hispanic Other<sup>1</sup> | 2/48 (4) |
| Not reported | 3/48 (6) |
| **Birth Information – Median (IQR)** |  |
| Gestational age, weeks | 38.7 (36.1, 39.2) |
| Birth weight, grams | 2570 (2215, 3140) |
| Birth length, cm | 48.3 (44.8, 49.6) |
| Head circumference, cm | 31.5 (30.0, 33.5) |
| **CMV Test Specimen – n/N (%)** |  |
| Saliva and urine | 20/48 (42) |
| Urine only | 15/48 (31) |
| Urine and Blood | 12/48 (25) |
| Saliva and Blood | 1/48 (2) |
| **Newborn Hearing Screening<sup>2</sup> – n/N (%)** |  |
| Passed – n/total tested (%) | 20/45 (44) |
| Failed – n/total tested (%) | 25/45 (56) |
| **Clinical Findings<sup>3</sup> – n/N (%)** |  |
| SGA/IUGR<sup>4</sup> | 24/48 (50) |
| Abnormal neuroimaging<sup>5</sup> | 23/48 (48) |
| Thrombocytopenia | 18/48 (38) |
| SNHL | 16/48 (33) |
| Premature | 14/48 (29) |
| Microcephaly | 14/48 (29) |
| Hepatitis | 11/48 (23) |
| Abnormal muscle tone | 10/48 (21) |
| Petechiae | 8/48 (17) |
| Hepatosplenomegaly | 4/48 (8) |
| Chorioretinitis | 3/48 (6) |
| Other<sup>6</sup> | 5/48 (10) |
| **Antiviral Treatment for cCMV – n/N (%)** |  |
| Prescribed antivirals | 32/48 (67) |
| Antiviral course completed | 25/32 (78) |
| Antiviral toxicity – n/total treated (%) | 11/32 (34) |
| Neutropenia | 9/11 (82) |
| Anemia | 2/11 (18) |
| Hepatitis | 2/11 (18) |

Data are presented as no./Total (%) for categorical measures and median (IQR) for continuous measures.
<sup>1</sup> Other: Asian (1) + “Race: I do not see my Race listed here; Ethnicity: Not Hispanic or Latino” (2)
<sup>2</sup> Total excluded (3): deceased - not tested (2), or missing newborn hearing screening (1)
<sup>3</sup> See Table S1 for study definitions.
<sup>4</sup> SGA: Small for Gestational Age; IUGR: Intrauterine Growth Restriction
<sup>5</sup> Cysts (16), Hemorrhage (8), Lenticulostriate vasculopathy (5), Calcification (4), Ventriculomegaly (2)
<sup>6</sup> Other findings: CMV pneumonitis (2), Optic nerve hypoplasia (1), Cholestasis (1), Coloboma of macula (1)

Follow-up beyond 12 months was available for 41/48 (85%) children, with a median follow-up duration of 74 months (IQR, 39–88 months). Nine children (22%) had no sequelae attributable to cCMV. Developmental delays occurred in 25/41 (61%), most often involving speech/language (16/25, 64%) or motor skills (15/25, 60%), with global developmental delay in 7/25 (28%) (Table 2). Among the 16 children with SNHL at birth, hearing worsened in 13 (81%), remained stable in 2 (13%), and improved in 1 (6%) (Figure 3). At the end of the follow-up period, 18/41 (44%) had SNHL, including two who developed late-onset hearing loss. All children with SNHL (18/18, 100%) required hearing aids or cochlear implants.

**Figure 2.**
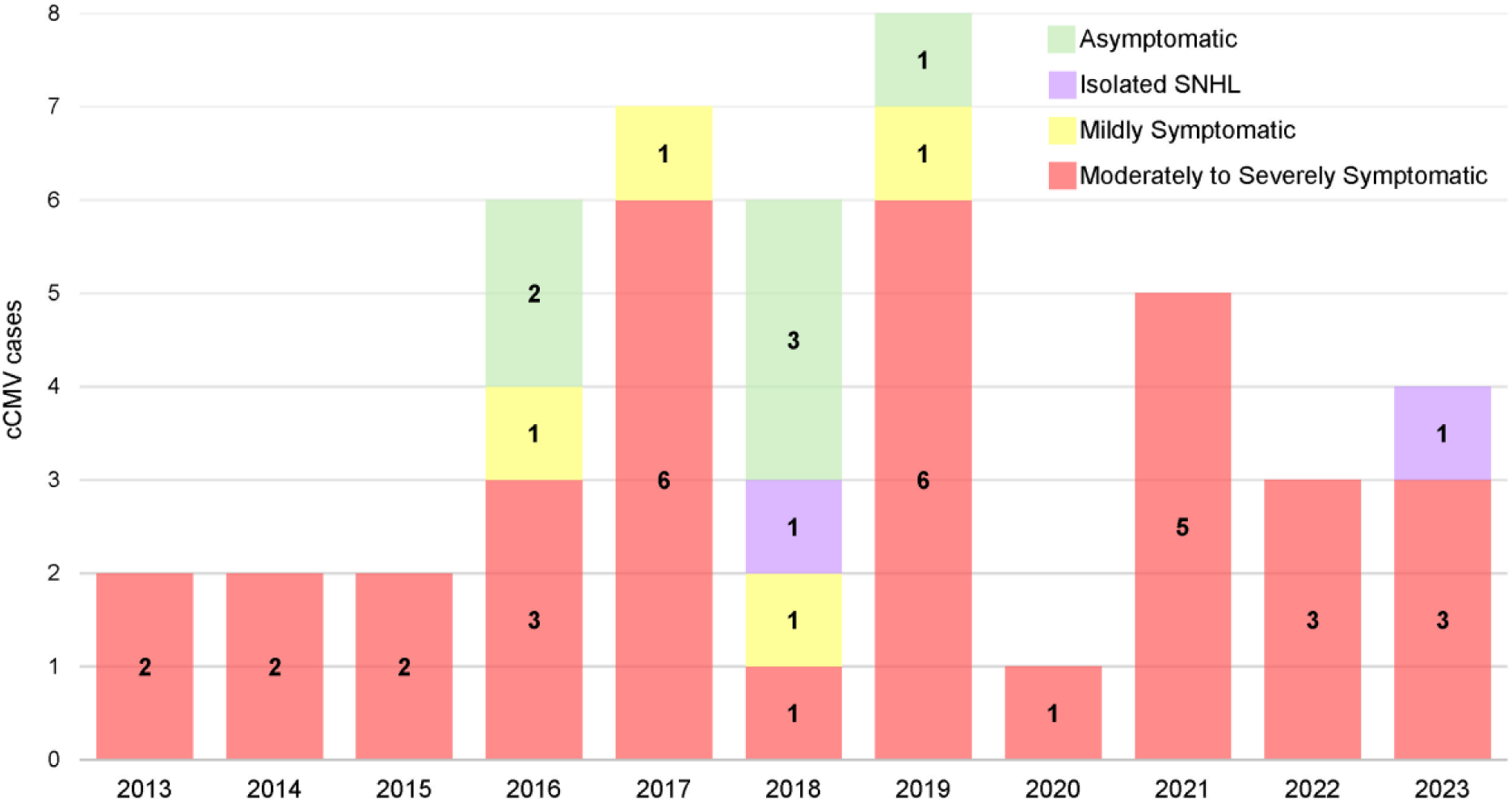
cCMV Disease Severity for each year. Annual distribution of congenital cytomegalovirus (cCMV) disease severity among confirmed cases, 2013–2023, with colors denoting severity: green (asymptomatic), purple (isolated sensorineural hearing loss), yellow (mildly symptomatic), and red (moderately to severely symptomatic).

**Figure 3.**
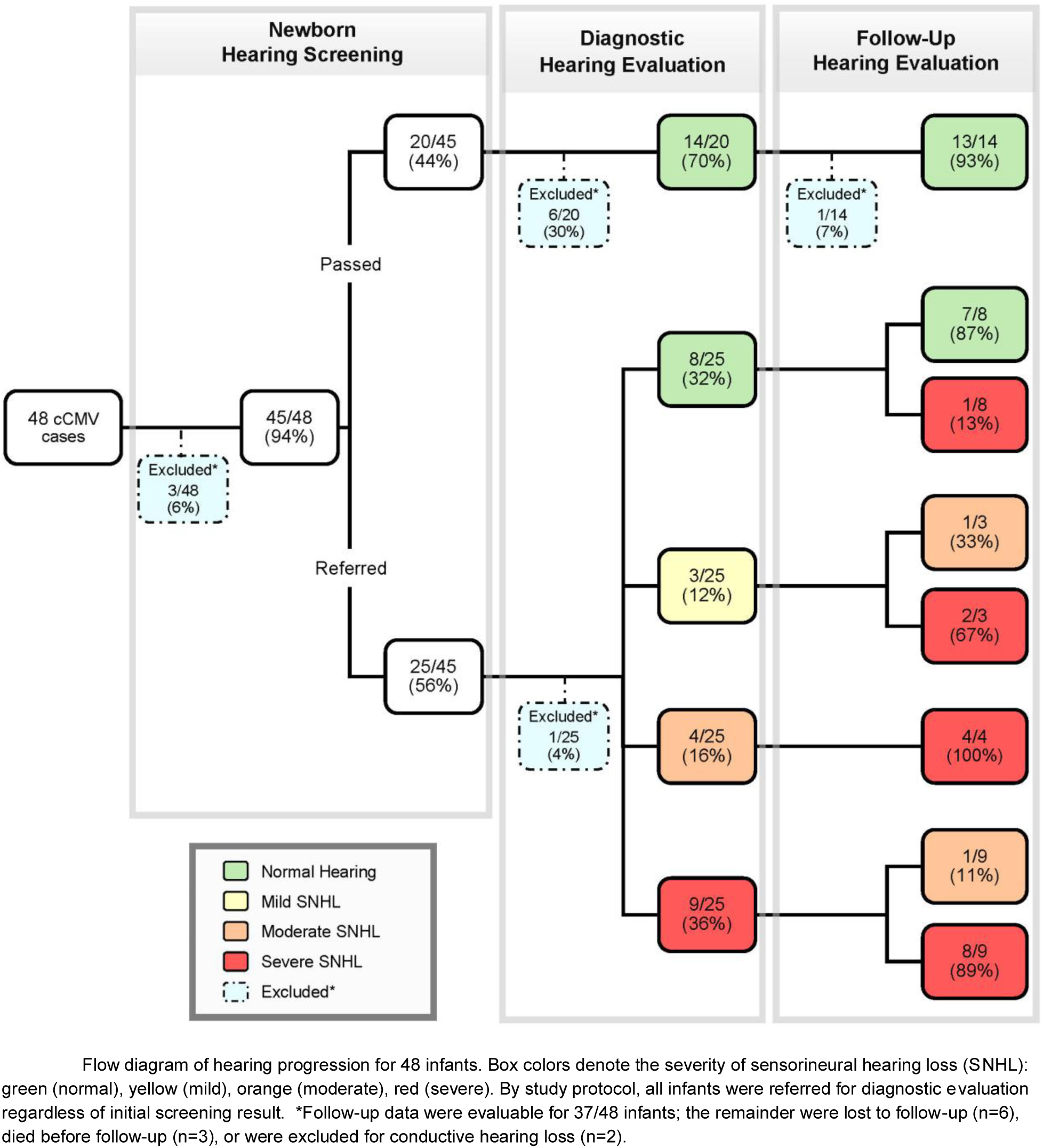
Progression of hearing status in 48 infants with congenital cytomegalovirus (cCMV) Progression of hearing status in 48 infants with cCMV, with color-coded categories for SNHL severity (green, normal; yellow, mild; orange, moderate; red, severe); follow-up was available for 37 infants, while 11 were lost, deceased, or excluded.

### Impact of Targeted Screening on Detection Rates

The estimated effects of the 2016 targeted screening law on annual cCMV diagnosis rates are shown in Figure 4. Prior to implementation of Connecticut’s 2016 hearing-targeted screening law, the mean annual incidence of cCMV was 1.08 per 10,000 births (95% CI, 0.22–1.95) with no significant change year-to-year (IRR, 1.00; 95% CI, 0.60–1.66; p = 1.00). After the 2016 law, there was an immediate and statistically significant increase in the rate of cCMV (IRR 3.58; 95% CI, 1.17-11.0; p = 0.026). The trend of new diagnoses remained stable from 2016 to 2019, averaging 4.09 new diagnoses per 10,000 births/year (95% CI, 2.60-5.58). In 2020, rates dropped relative to the post-law mean (IRR 0.37; 95% CI, 0.20-0.66; p = 0.001). Accounting for this transient decrease, the average incidence of diagnosed cCMV across the entire post-policy period was 2.96 per 10,000 births (95% CI: 2.07-3.86). The spectrum of disease severity also shifted after implementation of targeted screening (Figure 2). Prior to 2016, all detected cases were moderately to severely symptomatic at birth. In the post-policy period, 11 infants with clinically inapparent cCMV were identified. After a full diagnostic evaluation and long-term follow-up, 8 of these 11 infants (73%) were found to have either moderately-to-severely symptomatic disease or significant long-term sequelae, such as SNHL or developmental delay.

**Figure 4.**
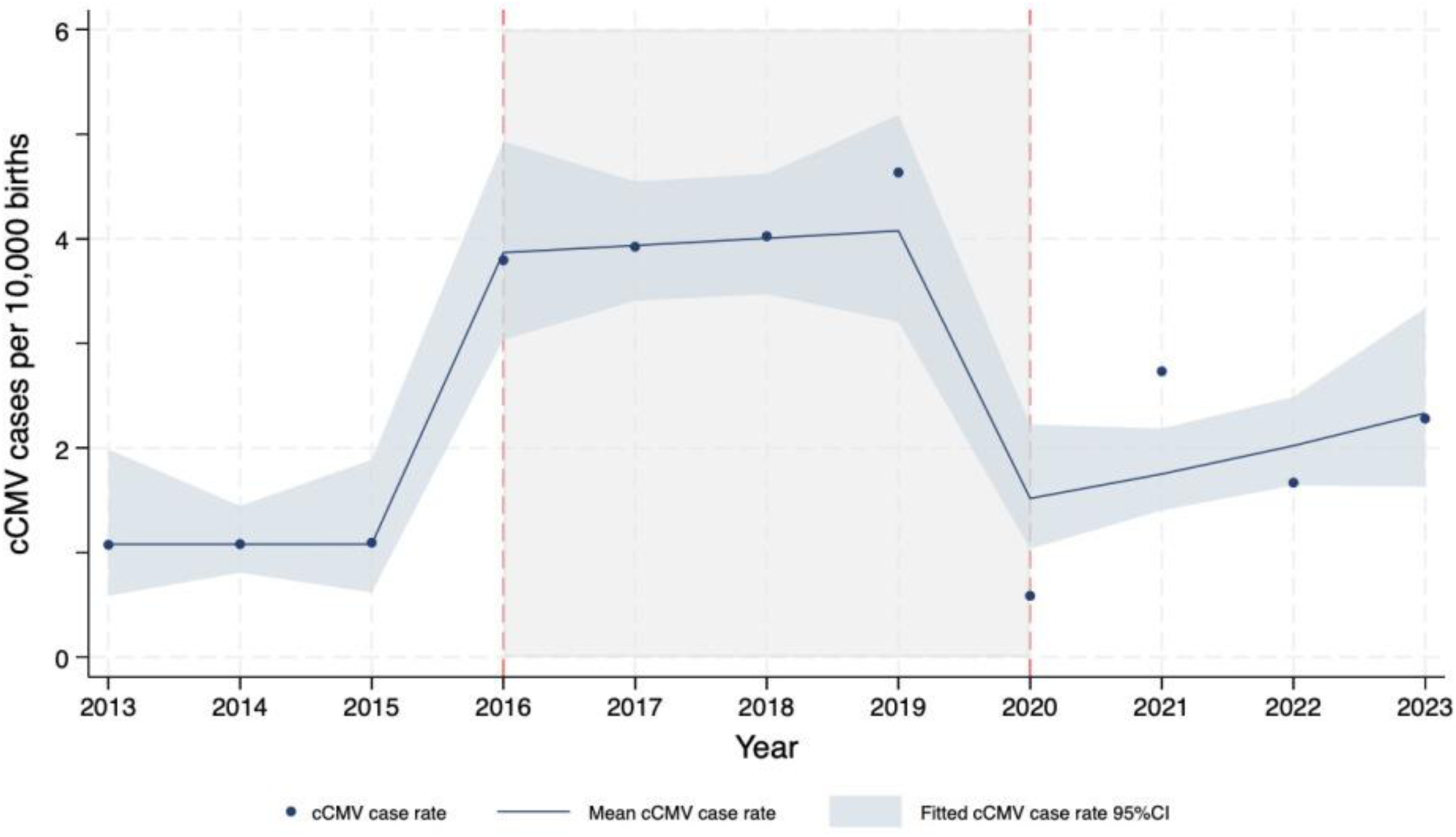
Observed and Predicted cCMV Case Rates per 10,000 births in Connecticut During the Period 2013 – 2023.

## Discussion

In this 11-year interrupted time series analysis, we found that Connecticut’s 2016 legislative mandate for hearing-targeted cCMV screening was associated with a 3.6-fold increase in the rate of cCMV diagnosis within a large healthcare network. This finding provides real-world evidence that such policies can substantially improve case ascertainment compared to reliance on clinical suspicion alone. Consistent with prior studies, the increase in diagnoses was largely driven by detection of clinically inapparent cases^23^. Importantly, “inapparent” did not imply benign, as 73% of these infants were later found to have clinically significant disease, including progressive hearing loss or developmental delays.^24^ This suggests that the hearing-targeted screening policy was effective at unmasking clinically significant disease.

However, gains in ascertainment did not consistently translate into improved hearing outcomes. In our cohort, hearing trajectories were largely unfavorable. Even though 88% of infants with SNHL at birth received antiviral therapy, a vast majority of them (81%, 13/16) experienced progression of hearing loss, with only one patient showing improvement. This observation complicates the assumptions of many cost-effectiveness models, which rely on efficacy data from idealized clinical trials and may therefore overestimate the real-world benefits of antiviral therapy on hearing outcomes.^25^ Although early diagnosis remains critical, its principal value in cCMV may lie less in reversing established injury and more in enabling timely audiologic interventions (e.g., hearing aids, cochlear implantation) that support language and cognitive development.^26,27^ Whether targeted screening meaningfully improves these long-term outcomes requires further dedicated study.

From a policy perspective, the choice between targeted and universal cCMV screening reflects a balance between maximizing case detection and resource allocation. Targeted screening is often favored for its cost-effectiveness, because it focuses on infants who fail the newborn hearing screen, a group with ∼4-fold higher prevalence of cCMV.^18,28^ However, its main disadvantage is its limited scope. Even at its peak, the post-policy incidence of cCMV in our system was 4.09 per 10,000 births, well below the estimated birth prevalence of 50–70 per 10,000 reported in universal screening studies.^1,2^ Targeted approaches miss late-onset SNHL, as well as non-audiologic symptoms or children that are entirely asymptomatic at birth.^29,30^ These missed cases include children who later developed clinically significant outcomes. This demonstrates that a hearing-targeted approach alone is insufficient and provides rationale for Connecticut’s recent transition to universal screening in 2025.

This study has several strengths. The quasi-experimental interrupted time series design, which uses pre-intervention data as a control, strengthens internal validity. Our cCMV detection rate prior to policy implementation (1.08 per 10,000 live births) is consistent with estimates from other U.S. studies that relied on clinical recognition and routine care, which have typically reported prevalence of ∼1–2 cases per 10,000 births.^31–33^ This suggests that our baseline data are reflecting the typical patterns of detection that arise from clinical recognition and routine clinical care. Another key strength was in the use of a large, longitudinal birth cohort with over a decade of observations. The systematic collection of clinical, neuroimaging, and longitudinal outcome data enabled precise estimation of cCMV incidence and characterization of disease progression within this population.

The study also has important limitations. First, as an observational study, causal inference is inherently constrained by the potential for residual confounding. Although the models adjusted for secular trends in birth rates and attempted to isolate the policy effect from contemporaneous influences such as the COVID-19 pandemic, unmeasured time-varying confounders may still have influenced the observed outcomes. Second, the study was conducted within a single healthcare network, which may limit external validity and generalizability to other healthcare systems or populations with differing screening practices or demographic characteristics. Third, variability in adherence to follow-up recommendations and incomplete outcome data for infants receiving care outside the network may have introduced outcome misclassification. However, the YNHHS’s participation in a statewide Health Information Exchange, which integrates data from all acute care hospitals and approximately half of ambulatory providers in Connecticut, partially mitigates this concern. Lastly, we did not evaluate the program’s cost-effectiveness, its psychological impact on families, or its comparative performance relative to universal screening strategies. These are important questions that require future research.

In conclusion, Connecticut’s hearing-targeted newborn cCMV screening mandate was associated with a substantial increase in case detection relative to the pre-policy period. However, this screening approach likely captured only a fraction of the disease burden. This real-world assessment of benefits and limitations can inform states and health systems as they design or refine cCMV screening strategies, including whether to adopt universal screening.

## Data Availability

All data produced in the present study are available upon reasonable request to the authors

---

**Figure S1.** Geographic distribution of study population. Connecticut map showing the average birth rates of towns served by Yale New Haven Health System (YNHHS) and affiliated hospitals, along with their locations.

**Figure S2.** Algorithm for cytomegalovirus (CMV) testing and follow-up. Clinical workflow for diagnostic testing and follow-up of infants who failed newborn hearing screening prior to hospital discharge.

## References

1. Casteels A, Naessens A, Gordts F, De Catte L, Bougatef A, Foulon W. Neonatal screening for congenital cytomegalovirus infections. J Perinat Med. 1999;27(2):116–21. doi:10.1515/jpm.1999.015

2. Dollard SC, Grosse SD, Ross DS. New estimates of the prevalence of neurological and sensory sequelae and mortality associated with congenital cytomegalovirus infection. Rev Med Virol. Sep–Oct 2007;17(5):355–63. doi:10.1002/rmv.544

3. Institute of Medicine Committee to Study Priorities for Vaccine D. The National Academies Collection: Reports funded by National Institutes of Health. In: Stratton KR, Durch JS, Lawrence RS, eds. Vaccines for the 21st Century: A Tool for Decisionmaking. National Academies Press (US) Copyright 2000 by the National Academy of Sciences. All rights reserved.; 2000.

4. Schleiss MR, Permar SR, Plotkin SA. Progress toward Development of a Vaccine against Congenital Cytomegalovirus Infection. Clin Vaccine Immunol. Dec 2017;24(12)doi:10.1128/cvi.00268-17

5. Pass RF, Arav-Boger R. Maternal and fetal cytomegalovirus infection: diagnosis, management, and prevention. F1000Res. 2018;7:255. doi:10.12688/f1000research.12517.1

6. Cannon MJ. Congenital cytomegalovirus (CMV) epidemiology and awareness. J Clin Virol. Dec 2009;46 Suppl 4:S6–10. doi:10.1016/j.jcv.2009.09.002

7. Dahle AJ, Fowler KB, Wright JD, Boppana SB, Britt WJ, Pass RF. Longitudinal investigation of hearing disorders in children with congenital cytomegalovirus. J Am Acad Audiol. May 2000;11(5):283–90.

8. Goderis J, De Leenheer E, Smets K, Van Hoecke H, Keymeulen A, Dhooge I. Hearing loss and congenital CMV infection: a systematic review. Pediatrics. Nov 2014;134(5):972–82. doi:10.1542/peds.2014-1173

9. Pinninti SG, Britt WJ, Boppana SB. Auditory and Vestibular Involvement in Congenital Cytomegalovirus Infection. Pathogens. Nov 20 2024;13(11)doi:10.3390/pathogens13111019

10. Lanzieri TM, Chung W, Flores M, et al. Hearing Loss in Children With Asymptomatic Congenital Cytomegalovirus Infection. Pediatrics. Mar 2017;139(3)doi:10.1542/peds.2016-2610

11. Kimberlin DW, Jester PM, Sánchez PJ, et al. Valganciclovir for symptomatic congenital cytomegalovirus disease. N Engl J Med. Mar 5 2015;372(10):933–43. doi:10.1056/NEJMoa1404599

12. Swanson EC, Schleiss MR. Congenital cytomegalovirus infection: new prospects for prevention and therapy. Pediatr Clin North Am. Apr 2013;60(2):335–49. doi:10.1016/j.pcl.2012.12.008

13. Rawlinson WD, Boppana SB, Fowler KB, et al. Congenital cytomegalovirus infection in pregnancy and the neonate: consensus recommendations for prevention, diagnosis, and therapy. The Lancet Infectious Diseases. 2017/06/01/ 2017;17(6):e177–e188. 10.1016/S1473-3099(17)30143-3

14. Leruez-Ville M, Foulon I, Pass R, Ville Y. Cytomegalovirus infection during pregnancy: state of the science. Am J Obstet Gynecol. Sep 2020;223(3):330–349. doi:10.1016/j.ajog.2020.02.018

15. Sorichetti B, Goshen O, Pauwels J, et al. Symptomatic Congenital Cytomegalovirus Infection Is Underdiagnosed in British Columbia. J Pediatr. Feb 2016;169:316–7. doi:10.1016/j.jpeds.2015.10.069

16. Wilson KL, Shah K, Pesch MH. Inconsistent Provider Testing Practices for Congenital Cytomegalovirus: Missed Diagnoses and Missed Opportunities. Int J Neonatal Screen. Nov 14 2022;8(4)doi:10.3390/ijns8040060

17. Marsico C, Kimberlin DW. Congenital Cytomegalovirus infection: advances and challenges in diagnosis, prevention and treatment. Ital J Pediatr. Apr 17 2017;43(1):38. doi:10.1186/s13052-017-0358-8

18. Gievers LL, Holmes AV, Loyal J, et al. Ethical and Public Health Implications of Targeted Screening for Congenital Cytomegalovirus. Pediatrics. Jul 2020;146(1)doi:10.1542/peds.2020-0617

19. Health CDoP. Congenital cytomegalovirus (cCMV) screening and follow-up. CT.gov. Accessed July 23, 2025. https://portal.ct.gov/DPH/Family-Health/EHDI/CMV

20. Haller T, Shoup A, Park AH. Should hearing targeted screening for congenital cytomegalovirus infection Be implemented? Int J Pediatr Otorhinolaryngol. Jul 2020;134:110055. doi:10.1016/j.ijporl.2020.110055

21. Committee on Infectious Diseases AAoP. Cytomegalovirus Infection. In: Kimberlin DW, Banerjee R, Barnett ED, Lynfield R, Sawyer MH, eds. Red Book: 2024–2027 Report of the Committee on Infectious Diseases. American Academy of Pediatrics; 2024:0.

22. Lanzieri TM, Caviness AC, Blum P, Demmler-Harrison G. Progressive, Long-Term Hearing Loss in Congenital CMV Disease After Ganciclovir Therapy. J Pediatric Infect Dis Soc. Jan 27 2022;11(1):16–23. doi:10.1093/jpids/piab095

23. Ronner EA, Glovsky CK, Herrmann BS, Woythaler MA, Pasternack MS, Cohen MS. Congenital Cytomegalovirus Targeted Screening Implementation and Outcomes: A Retrospective Chart Review. Otolaryngol Head Neck Surg. Jul 2022;167(1):178–182. doi:10.1177/01945998211044125

24. J ACBMEDHALJLNMAMTRJOCSST. Report on the Findings of the Universal cCMV Working Group. Vol. 2025. 2025. January. https://portal.ct.gov/dph/family-health/ehdi/cmv

25. Gantt S, Dionne F, Kozak FK, et al. Cost-effectiveness of Universal and Targeted Newborn Screening for Congenital Cytomegalovirus Infection. JAMA Pediatr. Dec 1 2016;170(12):1173–1180. doi:10.1001/jamapediatrics.2016.2016

26. Hirve R, Adams C, Kelly CB, et al. Effect of early childhood development interventions delivered by healthcare providers to improve cognitive outcomes in children at 0-36 months: a systematic review and meta-analysis. Arch Dis Child. Apr 2023;108(4):247–257. doi:10.1136/archdischild-2022-324506

27. Pesch MH, Lauer CS, Weinberg JB. Neurodevelopmental outcomes of children with congenital cytomegalovirus: a systematic scoping review. Pediatr Res. Jan 2024;95(2):418–435. doi:10.1038/s41390-023-02639-6

28. Kenneson A, Cannon MJ. Review and meta-analysis of the epidemiology of congenital cytomegalovirus (CMV) infection. Rev Med Virol. Jul–Aug 2007;17(4):253–76. doi:10.1002/rmv.535

29. Leruez-Ville M, Guilleminot T, Stirnemann J, et al. Quantifying the Burden of Congenital Cytomegalovirus Infection With Long-term Sequelae in Subsequent Pregnancies of Women Seronegative at Their First Pregnancy. Clin Infect Dis. Oct 23 2020;71(7):1598–1603. doi:10.1093/cid/ciz1067

30. Williams EJ, Kadambari S, Berrington JE, et al. Feasibility and acceptability of targeted screening for congenital CMV-related hearing loss. Arch Dis Child Fetal Neonatal Ed. May 2014;99(3):F230–6. doi:10.1136/archdischild-2013-305276

31. Lopez AS, Ortega-Sanchez IR, Bialek SR. Congenital cytomegalovirus-related hospitalizations in infants <1 year of age, United States, 1997-2009. Pediatr Infect Dis J. Nov 2014;33(11):1119–23. doi:10.1097/inf.0000000000000421

32. Messinger CJ, Lipsitch M, Bateman BT, et al. Association Between Congenital Cytomegalovirus and the Prevalence at Birth of Microcephaly in the United States. JAMA Pediatr. Dec 1 2020;174(12):1159–1167. doi:10.1001/jamapediatrics.2020.3009

33. G QESLEJMMC. Case-Finding and Validation of Congenital Cytomegalovirus (cCMV) Related Hospital Admissions in Michigan Infants under One Year of Age. 2016. October 2016. https://www.michigan.gov/-/media/Project/Websites/mdhhs/Folder1/Folder94/cCMV_Report_Final.pdf?rev=c6073d3d277e4b17830709881e01692c

