## Supplementary material for "Impact of a Statewide Hearing-Targeted Congenital Cytomegalovirus Screening Mandate: An 11-Year Interrupted Time Series Analysis": Table S1

### Supplementary Table

**Table S1**: Variable definition table along with cCMV diagnostic codes

| Category | Finding/Abnormality | Description/Criteria |
|---|---|---|
| **cCMV Diagnostic Codes** | ICD-9/10 codes for congenital CMV infection or CMV disease:<br>• 771.1, P35.1, 078.5; B25.x<br>SNOMED-CT codes for:<br>• Congenital cytomegalovirus infection (59527008)<br>• Cytomegalovirus infection (28944009)<br>• Cytomegalovirus (407444007)<br>• Cytomegalovirus hepatitis (186698009)<br>• Cytomegaloviral retinitis (22455005)<br>• Cytomegaloviral colitis (235749000), or<br>• Cytomegaloviral pneumonia (7678002) | |
| **Physical Examination: Clinical Findings** | Prematurity | Infants born at <37 weeks gestational age (GA). |
| | Small for Gestational Age (SGA) | Birth weight <10th percentile for GA. |
| | Microcephaly | Occipitofrontal circumference of <3rd percentile for GA. |
| | Neurological Abnormalities | Central or truncal hypotonia, hypertonia, or seizures. |
| | Ophthalmologic Abnormalities | Chorioretinitis, retinal scarring, or optic nerve hypoplasia. |
| **Structural Brain Abnormalities** | Intracranial calcifications, cysts, lenticulostriate vasculopathy, or hemorrhages on neonatal ultrasound, CT, or MRI. | |
| **Hematologic and Biochemical Abnormalities** | Anemia | Hematocrit <40% at 0-7 days of age, <35% at 8-14 days, and <30% at 15-28 days. |
| | Hepatitis | Jaundice and elevated direct bilirubin (>2.0 mg/dL) or elevated transaminases (≥40 U/mL). |
| | Neutropenia | Absolute neutrophil count of <1000 cells/$mm^3$. |
| | Thrombocytopenia | Platelet count of <150,000/$mm^3$. |

| Category | Finding/Abnormality | Description/Criteria |
| --- | --- | --- |
| **Hearing Evaluation** | Sensorineural hearing loss (SNHL) | Uni- or bilateral, assessed using Auditory Brainstem Response (ABR) testing or average hearing threshold across 0.5, 1, 2, and 4 kHz frequencies. Hearing status was classified based on thresholds in the more severely affected ear, as follows:<br>• Normal Hearing: 0-20 dB<br>• Mild Hearing Loss: 21-40 dB<br>• Moderate Hearing Loss: 41-70 dB<br>• Severe Hearing Loss: >70 dB |
| **Long-Term Neurodevelopmental Sequelae** | Developmental Delays and Neurobehavioral Disorders | Global developmental delay (≥2 domains affected), cognitive delay, speech and language delay, fine and/or gross motor delay, unspecified developmental delay, Autism Spectrum Disorder (ASD), Attention-Deficit/ Hyperactivity Disorder (ADHD). |
| | Other Neurologic Impairments | Cerebral palsy, seizure disorders, persisting abnormal muscle tone (hypotonia or hypertonia). |
| **cCMV Disease Severity**[1]* | Asymptomatic | No clinically apparent signs of cCMV disease and normal hearing. |
| | Isolated SNHL | No clinically apparent signs of cCMV disease, but presence of SNHL. |
| | Mildly Symptomatic | Presence of two or fewer transient (<2 weeks) or clinically insignificant findings, such as petechiae, mild hepatomegaly, thrombocytopenia, or mildly elevated alanine aminotransferase levels. |
| | Moderately to Severely Symptomatic | Presence of any of the following: a single severe or multiorgan disease (e.g., marked hepatosplenomegaly or significant liver enzyme abnormalities); life-threatening organ dysfunction; multiple persistent (≥2 weeks) CMV-related findings (e.g., thrombocytopenia, petechiae, hepatitis); central nervous system involvement (e.g., microcephaly, neuroimaging abnormalities, abnormal cerebrospinal fluid indices, |

| Category | Finding/Abnormality | Description/Criteria |
| --- | --- | --- |
| | | chorioretinitis, or CMV DNA detected in CSF); or more than two mild manifestations. |
| **cCMV Visible Symptoms** | Clinically Apparent | Documentation of at least one exam finding consistent with congenital infection, including small for gestational age, prematurity, microcephaly, abnormal tone, hepatomegaly, splenomegaly, petechiae, purpura, or seizures |
| | Clinically Inapparent | Infants with no visible cCMV symptoms as described above. |

* Adapted from Red Book


1. Committee on Infectious Diseases AAoP. Cytomegalovirus Infection. In: Kimberlin DW, Banerjee R, Barnett ED, Lynfield R, Sawyer MH, eds. Red Book: 2024–2027 Report of the Committee on Infectious Diseases: American Academy of Pediatrics; 2024:0.
