## Supplementary material for "Impact of a Statewide Hearing-Targeted Congenital Cytomegalovirus Screening Mandate: An 11-Year Interrupted Time Series Analysis": Figure S1

### Supplementary Figures

**Figure S1:** Connecticut map showing the average birth rates of the towns served by YNHHS and affiliated hospitals, and their locations

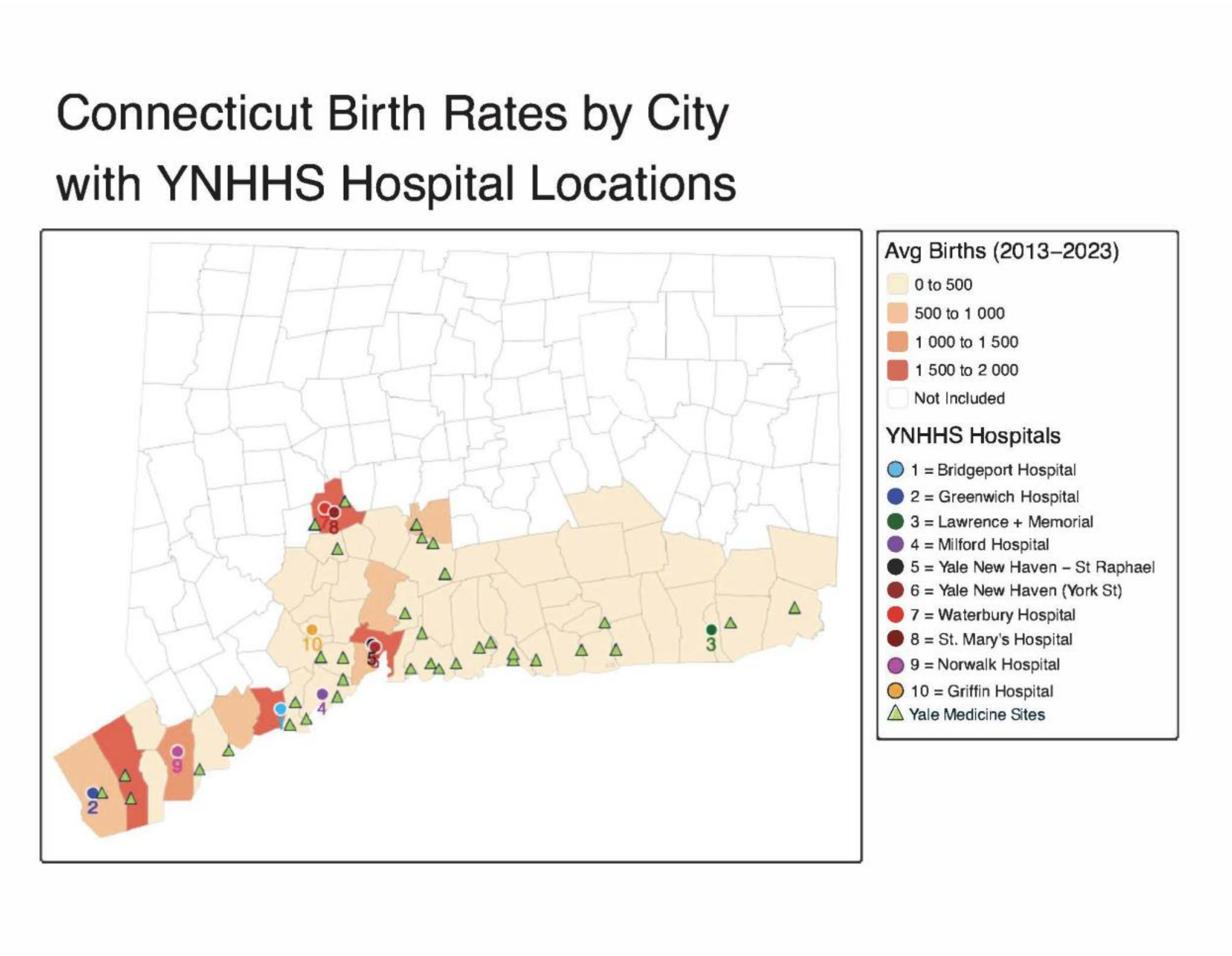
