## Supplementary material for "Impact of a Statewide Hearing-Targeted Congenital Cytomegalovirus Screening Mandate: An 11-Year Interrupted Time Series Analysis": Figure S2

### Algorithm for CMV testing and follow-up on infants who fails hearing screen prior to hospital discharge

Infant fails initial and repeat hearing screens (ABR)

Can urine be collected?

**Yes** → Send Urine for CMV PCR

- Urine CMV PCR is POSITIVE
  - Notify family and PCP of results.
  - Consult Pediatric ID and schedule appointment in ID clinic.
- Urine CMV PCR is NEGATIVE

**No** → Send Saliva for CMV PCR

- Saliva CMV PCR is NEGATIVE
- Saliva CMV PCR is POSITIVE
  1) Notify family and PCP of results.
  2) Send Urine for CMV PCR.
  3) **If urine CMV PCR POSITIVE:** Consult Pediatric ID.

Urine CMV PCR is NEGATIVE / Saliva CMV PCR is NEGATIVE:

- Notify family and PCP of results.
- No further CMV testing.
- Refer for formal audiology evaluation in 2-4 weeks.

Pediatric ID clinic evaluation

1) **Exam Findings**: Microcephaly; IUGR; Hepatosplenomegaly; Petechiae; Hypotonia.
2) **Head Ultrasound**: Periventricular calcifications; Cerebellar Hypoplasia; Ventriculomegaly.
3) **Laboratory Evaluation**: Anemia; Neutropenia; Thrombocytopenia; Transaminitis; Hyperbilirubinemia; BUN/Creatinine; CMV blood PCR.
4) **Ophthalmological Examination**: Chorioretinitis; Retinal scar; Optic atrophy.

Is the infant <1 month of age and are any of the above findings present?

**Yes**

**Treatment Plan:**

- **Medication:** Valganciclovir 16 mg/kg BID x 6 months (if normal BUN/Cr)
- **Follow-up Tests:**
  - 1 week after initiation: CBC, LFT.
  - Monthly monitoring if stable.
  - CMV viral load at 3 and 6 months.
- **Other Care:**
  1) Neurodevelopmental screen.
  2) Refer to Birth-to-3 if hearing impaired.
  3) MRI Brain if abnormal head ultrasound.
  4) Hearing and Ophthalmology evaluation every q6months until 1 year.
  5) Provide CMV prevention education.

**Drug Toxicity Protocol:**

1) **Hold Treatment** if:
   - ANC<500
   - Platelets <50k
   - ALT >10x baseline
2) **Resume** when resolved:
   a. ANC >750
   b. Platelets >50k
   c. ALT at baseline
3) **If recurrence**: Reduce dose or stop

**No**

1) **Discuss with parents**:
   - Lack of data in this population.
   - Valganciclovir MAY prevent hearing loss progression.
   - Potential medication side effects.
2) **Offer treatment**

**Family desires treatment** → Treatment Plan

**Family refuses treatment**

- Neurodevelopmental Screen
- Hearing and Ophthalmology evaluation q6months until 1 year of age.
- Provide CMV prevention education.
